# Pilot study of a multifaceted nurse-led antimicrobial stewardship intervention in residential aged care

**DOI:** 10.1101/2023.10.31.23297824

**Authors:** Natali Jokanovic, Sue J Lee, Terry Haines, Sarah N Hilmer, Yun-Hee Jeon, Laura Travis, Darshini Ayton, Eliza Watson, Tess Tsindos, Andrew J Stewardson, Rhonda L Stuart, Allen C Cheng, Trisha N Peel, Anton Y Peleg, the START Trial Group

## Abstract

**Objective:** To evaluate the feasibility of a nurse-led antimicrobial stewardship (AMS) program in two Australian residential aged care homes (RACHs) to inform a stepped-wedged, cluster randomised controlled trial (SW-cRCT).

**Methods:** A mixed-methods pilot study of a nurse-led AMS program was performed in two RACHs in Victoria, Australia between July and December 2019. The AMS program comprised education, infection assessment and management guidelines, and documentation to support appropriate antimicrobial use in urinary, lower respiratory and skin/soft tissue infections. The program was implemented over three phases over five months: 1) pre-implementation education and integration (1-month); 2) implementation of the intervention (3-months); 3) post-intervention evaluation (1-month). Baseline RACH and resident data and weekly infection and antimicrobial usage was collected. Feedback on intervention resources and implementation barriers were identified from semi-structured interviews, online staff questionnaire and researcher field notes.

**Results:** Six key barriers to implementation of the intervention were identified and used to refine the intervention; aged care staffing and capacity, access to education, resistance to practice change, role of staff in AMS, leadership and ownership of the intervention at the RACH and organisation-level, and expectations from family. A total 61 antimicrobials were prescribed for 40 residents over the 3-month intervention period. Overall, 48% of antibiotics did not meet the minimum criteria for appropriate initiation (respiratory 73%; urinary: 54%; skin/soft tissue: 0%).

**Conclusions:** Several barriers and opportunities to improve the implementation of AMS in RACHs were identified. Findings were used to inform a revised intervention to be evaluated in a larger SW-cRCT.

## Introduction

Antimicrobial resistance (AMR) is a significant global concern and increasing within residential aged care homes (RACHs).^1, 2^ Driven by high rates of antibiotic use (up to 80% of residents),^3, 4^ which is often inappropriate,^4^ and the potential for AMR transmission across healthcare settings,^2, 5^ RACHs remain an important setting to target antimicrobial stewardship (AMS) initiatives.

AMS within hospitals is well-established with demonstrable reductions in antibiotic use,^6^ however their impact and key components within RACHs remains uncertain.^6, 7^ Interventions are frequently multifaceted and multidisciplinary, incorporating education, guidelines, and audit and feedback.^8^ RACHs are complex settings and present several challenges for successful implementation of AMS including staffing mix, workload, organisational structure, onsite availability of physicians and pharmacists and increasingly complex residents with multimorbidity and cognitive impairment.^9^ Recent randomised controlled trials have sought to investigate the impact of AMS on antibiotic use and AMR in RACHs,^10, 11^ however few have explored the barriers and facilitators of implementing AMS,^12^ particularly in Australian RACHs.^13, 14^

This study aimed to evaluate the feasibility of a nurse-led AMS intervention program in two Australian RACHs to inform a larger stepped-wedge, cluster randomised controlled trial (SW-cRCT).^11^

## Methods

### Study design and setting

A mixed-methods pilot study was performed in two RACHs in regional and metropolitan Victoria, Australia. Both RACHs provided 24-hour nursing care with general practitioner (GP) support. The study was performed over three phases between July-December 2019: 1) Pre-implementation (1-month education and intervention integration); 2) Implementation (3-months); 3) Post-intervention feedback (1-month).

### Intervention

Full intervention procedures are detailed in the published protocol.^11^ In brief, the nurse-led AMS intervention comprised education, RACH-specific guidelines, documentation forms and fact sheets to support appropriate antimicrobial prescribing for urinary tract infections (UTIs), lower respiratory tract infections (LRTIs), and skin and soft tissue infections (SSTIs). Education included face-to-face education, an online interactive workbook with equivalent information and fact sheets targeting improved diagnosis and antimicrobial management of common infections. RACH staff, GPs and pharmacists had access to all resources.

Education was provided by the study coordinator (research pharmacist) to RACH staff, including registered nurses (RNs), enrolled nurses (ENs), personal care attendants (PCAs) and clinical managers. Residents and families received monthly face-to-face education and a fact sheet.

RACH-specific guidelines were developed to support the initial assessment and antimicrobial management of UTIs, LRTIs and SSTIs, adapted from existing antibiotic initiation criteria,^15^ infection surveillance^16^ and prescribing guidelines.^17^ Assessment guidelines included minimum signs and symptoms of infection for antibiotic initiation, investigations and considerations for hospitalisation.

### Implementation

To support implementation, the nursing leadership team (clinical manager(s), general manager) were consulted prior and during the intervention to tailor implementation. The leadership team selected a “nurse champion” responsible for education completion, distribution and placement of resources onsite, and staff compliance with intervention procedures.

### Data collection

Baseline RACH, staff and resident data included occupancy, staffing mix and resident characteristics. All systemic antimicrobial use, infections and hospitalisations were collected from paper-based residents’ medical records. These data were collected to provide insight into baseline infection and antimicrobial rates as the short intervention period was insufficient to evaluate intervention effectiveness.

Post-intervention qualitative feedback included one-on-one semi-structured interviews and an online staff questionnaire covering an understanding of antibiotic appropriateness and resistance, and usability and usefulness of intervention resources. Interviews were audio-recorded and performed by two qualitative researchers (EW and TT). Researcher field notes documented observations related to intervention delivery.

### Analysis

Transcribed interviews and field notes were coded and thematically analysed independently by two researchers (NJ and LT) using inductive and deductive approaches to identify implementation barriers (NVivo, v20.3). Questionnaire data were summarised descriptively and considered alongside the initial themes to develop the final list of themes. Resident and antimicrobial data were summarised descriptively (Stata, v17.0).

### Ethics

Ethical approval was obtained from the Alfred Hospital Human Research Ethics Committee (HREC) (HREC/18/Alfred/591). Consent to obtain data from residents’ medication records was waived. Written informed consent were obtained for interviews. Consent was implied on completion of the online questionnaire.

## Results

### Resident and RACH characteristics

RACH-One comprised 60 beds serviced by up to 70 staff (15 RNs/ENs, 55 PCAs), 1 clinical manager and 16 GPs. RACH-Two comprised 75 beds serviced by up to 80 staff (20 RNs/ENs, 60 PCAs), 2 clinical managers and 10 GPs.

A total 135 residents were enrolled (Table 1). Residents were primarily female (71%), median 90 (IQR: 83-93) years of age, diagnosed with dementia (60%), required full assistance with activities of daily living (70%), and resided in their RACH for a median 31 (IQR: 10-59) months.

**Table 1.**
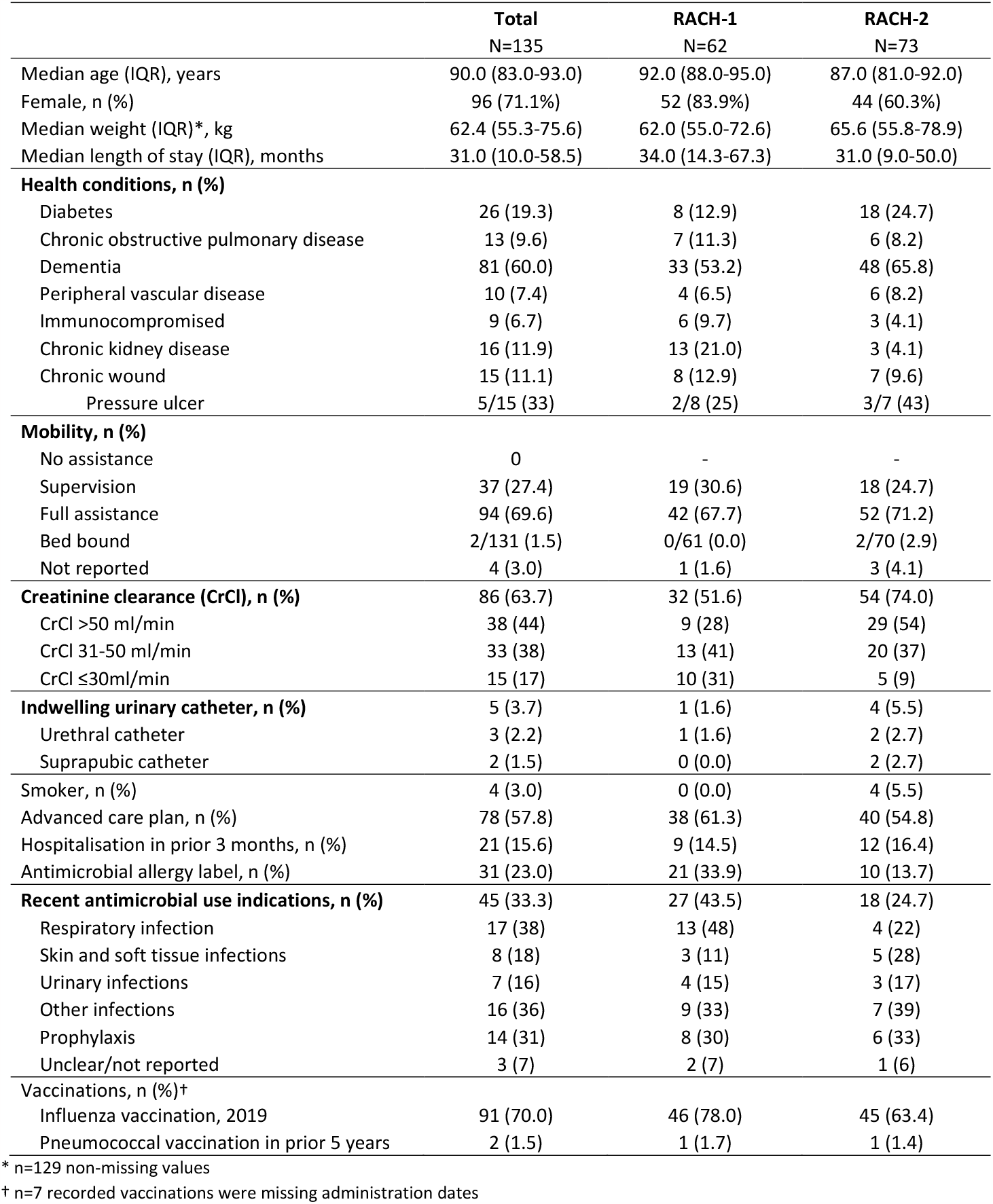
Resident baseline characteristics.

### Antimicrobial use

A total 124 suspected infections resulted in 61 antimicrobial prescriptions across 40 residents over three months (Supplementary Table S1). Up to 21% of residents were prescribed an antimicrobial on any given month (range: 2-21%). Antimicrobials were prescribed primarily for the treatment of UTIs (n=13, 21%), RTIs (n=22, 36%) and SSTIs (n=15, 25%). Half of antibiotic prescriptions (n=23/48, 48%) were inappropriate when assessed against minimum criteria (respiratory: 73%; urinary: 54%; skin/soft tissue: 0%).

### Implementation

Both RACHs selected a clinical manager, a RN by background, to provide study leadership onsite. Up to four face-to-face 1-hour and twice weekly 15-minute education at nursing handover meetings were provided at each RACH. Clinical managers prioritised the attendance of regularly rostered RNs/ENs for 1-hour education as they were perceived to have the greatest input in intervention procedures (attendance of total employed including casual workforce: 30% RNs/ENs, 30-35% PCAs).

Post-intervention feedback was obtained from seven one-on-one interviews (2 clinical managers, 3 RNs, 1 general manager, 1 resident) and online questionnaire (n=22). The questionnaire was completed by 4 RNs, 2 ENs, 13 PCAs, 2 clinical managers and 1 GP.

Overall, intervention resources were well-received. Of the questionnaire respondents, over 85% reported the resources were moderately-extremely useful in supporting improved antibiotic use and at least 77% were moderately-extremely likely to use them again. Improvements focused on simplifying education for PCAs and reducing content within guidelines and forms.

Six key themes related to implementation barriers identified from researcher field notes, interviews and the questionnaire included aged care staffing and capacity, education completion, resistance to practice change, staff roles in AMS, leadership and intervention ownership at the RACH and organisation-level, and family expectations (Table 2). Key improvements included strategies to increase education accessibility and completion (online, mandatory completion, staff renumeration), guideline integration into standard operating procedures, and increased engagement and capacity building of RACH and organisation-level leadership targeting uptake, compliance and intervention ownership.

**Table 2.** Barriers to implementation of antimicrobial stewardship and illustrative quotes from aged care staff* interviews (n=6), questionnaire (n=22) and researcher field notes.

| Themes | Subthemes | Summary from interviews, questionnaire and researcher field notes | Illustrative quotes from staff interviews |
| --- | --- | --- | --- |
| Aged care staffing and capacity | - Aged care staffing shortages<br>- High staff turnover<br>- Reliance on casual staffing<br>- Time management pressures<br>- Lack of confidence to challenge requests for antimicrobials | - RNs reported they were time-poor, overwhelmed with paperwork and were often short-staffed, resulting in reduced capacity to undertake education, provide adequate supervision of PCAs and integrate new procedures into their workflow.<br>- Reliance on casual staffing who are unfamiliar with new and existing procedures at the home.<br>- Clinical managers reported time management was often an issue and staff need to prioritise completing existing care over new initiatives and additional paperwork.<br>- Lack of confidence of aged care staff communicating and challenging family members' beliefs around antimicrobial use. | *"I'm run off my feet with lots of other things." – RN 3*<br>*"I guess time management...Resident stuff takes time. So some days it's really busy and there's days when it's you know stable...I think the main thing is trying to make sure the resident is safe and I don't know clinically they are looked after, because there's other things to be spending than going through paperwork I guess" – Clinical Manager 1* |
| Completion of education | - Insufficient time to complete education<br>- Lack of awareness to complete education<br>- Prioritisation of RNs and ENs over PCAs<br>- Mode of delivery<br>- Lack of mandatory education requirement<br>- Lack of reimbursement to complete education | - Pre-existing face-to-face education sessions preferentially target RNs and ENs, are not always mandatory and may not be paid if outside staff member's rostered hours.<br>- Clinical managers prioritised RN/EN attendance over PCAs, limiting the opportunity for upskilling of PCAs in AMS.<br>- Frequent short staffing and the absence of RN cover minimised attendance.<br>- Absence of payment for attendance to intervention education for staff who are not regularly rostered and unable to attend during working hours.<br>- Staff questionnaire: up to 60% prefer face-to-face education over online and/or written material. | *"Being the RN on the floor, unless they replace me, it's really, really hard to go off the floor for any length of time...so to actually get up to some of the lectures is just about impossible for me." – RN 3*<br>*"Yeah definitely if it's an at work thing we would definitely do it but I'm not really sure... if they directed it as mandatory we could do it. I mean for me I wasn't really told we have to do this but if I get told to then I would definitely go through with that." – RN 1* |
| Resistance to change | - Reliance on existing behaviours, knowledge and practices | - Aged care staff reported difficulty changing existing behaviours and practices towards infection assessment and testing. | *"[Reverting back to existing practices] It's just, it's what I've always known. You know, people are resistant to change and it's hard to adapt. It's not that I don't think it's a great tool, I think it absolutely is, it's just different. It's* |
|  | - Perception of lack of inappropriate antimicrobial use | - Difficulty changing long-standing procedures e.g. dipstick urine analysis/full ward tests in the absence of additional signs and symptoms beyond behaviour change.<br>- Aged care staff reported insufficient time to refer to intervention guidelines and instead relied on pre-existing clinical knowledge.<br>- Perception among aged care staff (all levels) that antimicrobial use was not necessarily a problem at their RACH. | *something different that I'm not used to." – Clinical Manager 2*<br>*"In terms of the guidance of it, has been helpful in some ways but there's always factors that will stop that from happening which includes staff will do a full ward test if they think that the resident's confused and they will continue to send it to the pathology to get it tested and also getting GPs involved in it as well would be a hard thing...and locums as well...like we can't stop them from starting antibiotics" – Clinical Manager 1*<br>*"I don't have time to actually look at it [resources] and I just use my basic knowledge, really nursing skills. I didn't really use these forms although I think it would be helpful for new nurses to look at so I still continue with what my nursing skills are. – Clinical Manager 1*<br>*"The knowledge is improving but I think it's more habit. I think its years and years and years of habit." – RN 3* |
| Role of staff in antimicrobial stewardship | - Difference in perceptions of role of PCAs<br>- Self-perceived lack of impact on prescribing by aged care staff | - Clinical managers and RNs felt role of PCAs was limited to identification of early symptoms, RNs as primary decision-makers in the assessment of suspected infections including testing and referral.<br>- Knowledge gap among PCAs prevents an active role within AMS.<br>- Staff survey: 62% (n=8/13) of PCAs agreed to strongly agreed to having an important role in AMS at their RACH.<br>- Aged care staff did not perceive they were well positioned to influence antimicrobial prescribing decisions and was the sole role of the GP.<br>- Aged care staff ambivalence over their individual role and impact on antimicrobial prescribing.<br>- Community pharmacists had a limited role outside of supply due to time constraints. | *"The PCAs here aren't involved in this stuff, they do ADLs and feed people and give them drinks and that's about all that goes mostly from what I can tell in this facility." – RN 3*<br>*"I don't think they [PCAs] really understand sometimes why we do stuff. They don't see the advantage of telling you someone was vomiting and it's like, this was Friday, it's like well why didn't you tell me? 'Oh it was only a little vomit' Well you report and I will decide if I follow it up or not. Subsequently the woman went to hospital, they didn't understand that I need to know so there's a gap there." – RN 3*<br>*"I mean definitely there are some people that are on antibiotics constantly that shouldn't be on them, but that's not our clinical judgement along with that, that is the doctors...at the end of the day I'm just a registered nurse, I don't have much leeway." – Clinical Manager 2*<br>*"It would probably be good as a 'we've tried doing this' kind of thing, we've tried implementing new ways of trying to improve infections and antibiotic use but in the*<br>*end it's really up to the doctors who prescribe these antibiotics and all that stuff."* – Clinical Manager 1 |
| Leadership and ownership at the RACH and organisation-level | - Lack of integration into standard operating procedures<br>- Duplicated procedures<br>- Insufficient awareness of new procedures<br>- Insufficient reinforcement to follow new procedures<br>- Competing pressures | - Lack of integration of intervention procedures into standard operating procedures and duplicated infection policies at the organisation-level.<br>- Staff duplicated notes across intervention forms, progress notes and incident reporting software.<br>- Insufficient awareness and reinforcement from designated onsite trial leaders/clinical managers to complete education and refer to intervention guidelines and forms.<br>- Competing pressures including concurrent changes in other procedures and staffing at the facilities taken priority over implementation of the intervention. | *"If [Aged Care Provider] says that this is what we have to do then I guess that's something that we need to implement...but because it's just a trial I think I'm sort of just like mmm you know, there's other changes that I've had to implement that are more important than this, so I haven't really encouraged the use of it I guess. There's too many changes that are happening at the moment that outweighs this."* – Clinical Manager 1<br>*"I mean if we get told 'we have to use this one' then definitely we can. I think it's just that not all of us were really aware about that...but yeah, I think more ongoing reinforcement that I'm using these [suspected infection] forms and all that would help us."* – RN 1<br>*"At this point what we do is write in the progress notes anyway, so filling in one of these [suspected infection forms] is repeating what you've already written so you sort of do it, I don't know, as a plus not necessarily for any particular reason it's just something extra you are doing on top of what you're already doing."* – RN 3 |
| Expectations from family | - Pressure to prescribe antimicrobials | - Pressure to prescribe antimicrobials from family members in the absence of appropriate indications for use. | *"Sometimes it's hard because we have family members that are insistent on their Mum or Dad having antibiotics, even though there is not really any clinical indication that they need it, like maybe the full ward test was clear or they were asymptomatic, but they are still quite insistent because they believe that Mum's coming down with a UTI or chest infection, so we have to advocate, just give them the information they need and get the GP on board as well."* – General Manager |

\*Aged care staff: registered and enrolled nurses, personal care assistants and clinical manager

## Discussion

This study evaluated the feasibility of implementing a nurse-led AMS program across two RACHs to inform a multifaceted AMS intervention for a SW-cRCT. Overall, the AMS program was well-received however several opportunities to improve implementation were identified. Implementation themes included staff perceptions of their role within AMS, and workforce and workload challenges,^13, 14, 18^ highlighting the complexities of implementing quality improvement programs within RACHs. Half of all antibiotics for UTIs, LRTIs and SSTIs did not meet minimum criteria for initiation, supporting the need for strategies to improve appropriate antimicrobial use.

Opportunities to improve implementation targeted education uptake and perceptions of RACH staff roles within AMS. PCAs comprise 70% of the Australian RACH workforce^19^ and are often the first to identify symptoms and provide information to residents and families. Despite this, clinical managers and RNs did not perceive a significant role for PCAs within AMS, citing key knowledge gaps, conflicting with PCAs self-perceived role and interest in improving their knowledge. This is consistent with previous Australian RACH staff interviews, identifying exclusion of PCAs from AMS education despite interest to improve antimicrobial use.^14^ Aged care staff did not perceive their role extended to influencing prescribing decisions, expressing it was the GP’s role and a lack of confidence in challenging decision-making, consistent with previous Australian aged care surveys.^20^ Addressing these knowledge and confidence gaps requires inclusive education and strategies to support attendance including flexible modes, and dedicated time and remuneration for completion.

Workforce and workflow challenges are consistent barriers to implementation of quality improvement programs.^14, 18^ Staff turnover, insufficient number of RNs and reliance on agency staff were identified as key barriers in this pilot and internationally.^14^ Aged care staff were uncertain how to prioritise new AMS procedures in the setting of competing pressures and organisational changes. Tailoring AMS workflow under the constraints of existing workforce challenges and prioritising AMS in RACHs is necessary to drive acceptance and adherence.

This study had several strengths and limitations. The intervention was championed at each RACH by existing clinical managers to support implementation. Regional and metropolitan sites provided broader representation of implementation barriers. Ongoing feedback during the intervention was provided informally by staff and contributed to revisions that informed the final intervention and implementation plan for a SW-cRCT. High staff turnover and poor availability of staff (particularly PCAs) at the time of evaluation limited our sample size. Although both RACHs were managed by the same provider, variability in culture, workforce and procedures may limit generalisability to other RACHs more broadly.

## Conclusions

This pilot identified several barriers and opportunities to improve implementation of AMS in RACHs. Findings were used to inform a revised intervention to be evaluated in a SW-cRCT across 12 RACHs.

## Data Availability

All data produced in the present study are available upon reasonable request to the authors

## Acknowledgements

The authors thank all staff and residents at participating RACHs for their support and involvement during this study.

## Collaborators

The START trial Principal Investigator: Professor Anton Peleg, The START co-investigators: Professor Terry Haines, Professor Allen Cheng, Professor Trisha Peel, Professor Kathryn Holt, Professor Sarah Hilmer, Professor Yun-Hee Jeon, Associate Professor Andrew Stewardson, Professor Rhonda Stuart. Statisticians: Dr Sue J Lee, Associate investigators: Dr Daniel Wilson, Associate Professor James Trauer, Professor Marilyn Cruickshank, Dr Nicola De Maio, Bupa Aged Care Australia, Bupa Health Foundation. START Trial Coordination and Administration: Dr Natali Jokanovic (START Clinical Trial Coordinator), Ms Janine Roney. Department of Infectious Diseases, Microbiology Laboratory: Dr Jessica Wisniewski.

## Transparency declarations

None to declare.

## References

1. Australian Commission on Safety and Quality in Heatlh Care. Antimicrobial stewardship in Australian Health Care. Sydney: ASCQHC; 2018.

2. Lim CJ, Cheng AC, Kennon J, et al. Prevalence of multidrug-resistant organisms and risk factors for carriage in long-term care facilities: a nested case-control study. J Antimicrob Chemother. 2014;69:1972–80.

3. Sluggett JK, Moldovan M, Lynn DJ, et al. National trends in antibiotic use in Australian residential aged care facilities, 2005-2016. Clin Infect Dis. 2020.

4. van Buul LW, van der Steen JT, Veenhuizen RB, et al. Antibiotic use and resistance in long term care facilities. J Am Med Dir Assoc. 2012;13:568.e1-13.

5. Mortensen E, Trivedi KK, Rosenberg J, et al. Multidrug-resistant Acinetobacter baumannii infection, colonization, and transmission related to a long-term care facility providing subacute care. Infect Control Hosp Epidemiol. 2014;35:406–11.

6. Zay Ya K, Win PTN, Bielicki J, et al. Association between antimicrobial stewardship programs and antibiotic use globally: a systematic review and meta-analysis. JAMA Netw Open. 2023;6:e2253806.

7. Aliyu S, Travers JL, Heimlich SL, et al. Antimicrobial Stewardship Interventions to Optimize Treatment of Infections in Nursing Home Residents: A Systematic Review and Meta-Analysis. J Appl Gerontol. 2022;41:892–901.

8. Nguyen HQ, Tunney MM, Hughes CM. Interventions to improve antimicrobial stewardship for older people in care homes: a systematic review. Drugs Aging. 2019;36:355–69.

9. Lim CJ, Kwong MW, Stuart RL, et al. Antibiotic prescribing practice in residential aged care facilities--health care providers’ perspectives. Med J Aust. 2014;201:98–102.

10. Mitchell SL, D’Agata EMC, Hanson LC, et al. The Trial to Reduce Antimicrobial Use in Nursing Home Residents With Alzheimer Disease and Other Dementias (TRAIN-AD): A Cluster Randomized Clinical Trial. JAMA Intern Med. 2021;181:1174–82.

11. Jokanovic N, Haines T, Cheng AC, et al. Multicentre stepped-wedge cluster randomised controlled trial of an antimicrobial stewardship programme in residential aged care: protocol for the START trial. BMJ open. 2021;11:e046142.

12. Baier RR, Jump RLP, Zhang T, et al. Feasibility of a nursing home antibiotic stewardship intervention. J Am Med Dir Assoc. 2022;23:1025–30.

13. Lim CJ, Kwong M, Stuart RL, et al. Antimicrobial stewardship in residential aged care facilities: need and readiness assessment. BMC Infect Dis. 2014;14:410.

14. Hall J, Hawkins O, Montgomery A, et al. Dismantling antibiotic infrastructures in residential aged care: The invisible work of antimicrobial stewardship (AMS). Soc Sci Med. 2022;305:115094.

15. Loeb M, Bentley DW, Bradley S, et al. Development of minimum criteria for the initiation of antibiotics in residents of long-term-care facilities: results of a consensus conference. Infect Control Hosp Epidemiol. 2001;22:120–4.

16. Stone ND, Ashraf MS, Calder J, et al. Surveillance definitions of infections in long-term care facilities: revisiting the McGeer criteria. Infect Control Hosp Epidemiol. 2012;33:965–77.

17. Antibiotics [published 2019] In: eTG complete [digital] [Internet]. Therapeutic Guidelines Limited; 2020.

18. Tappen RM, Wolf DG, Rahemi Z, et al. Barriers and facilitators to implementing a change initiative in long-term care using the INTERACT® quality improvement program. Health Care Manag (Frederick). 2017;36:219–30.

19. Mavromaras K, Knight G, Isherwood L, et al. The aged care workforce, 2016. Australian Government; 2017.

20. Lo SY, Reeve E, Page AT, et al. Attitudes to drug use in residential aged care facilities: a crosssectional survey of nurses and care staff. Drugs Aging. 2021;38:697–711.

