## Supplementary Table S1 for "Pilot study of a multifaceted nurse-led antimicrobial stewardship intervention in residential aged care"

**Table S1.** Antimicrobials prescribed over the three-month intervention period, by indication (n=61 prescriptions from 40 residents).

|  | **Total** | **RACH-1** | **RACH-2** |
| --- | --- | --- | --- |
| Residents prescribed an antimicrobial, n | 40 | 16 | 24 |
| Antimicrobial prescriptions, n | **61** | **26** | **35** |
| **Indications, n (%)*** |  |  |  |
| **Urinary tract infections, n (%)** | **13 (21)** | **6 (23)** | **7 (20)** |
| UTI | 7 | 5 | 2 |
| Catheter-associated UTI | 6 | 1 | 5 |
| **Respiratory tract infection, n (%)** | **22 (36)** | **6 (23)** | **16 (46)** |
| Nonspecific respiratory infection | 13 | 4 | 9 |
| Pneumonia | 5 | 1 | 4 |
| Other | 4 | 1 | 3 |
| **Skin/Soft tissue infections, n (%)** | **15 (25)** | **6 (23)** | **9 (26)** |
| Nonspecific wound | 2 | 1 | 1 |
| Cellulitis | 2 | 2 | 0 |
| Ulcer | 5 | 2 | 3 |
| Shingles | 2 | 0 | 2 |
| Other | 4 | 1 | 3 |
| **Prophylaxis, n (%)** | **2 (3)** | **1 (4)** | **1 (3)** |
| Post-surgery | 1 | 1 | 0 |
| Urinary tract infection | 1 | 0 | 1 |
| **Other infections, n (%)**† | **5 (8)** | **4 (15)** | **1 (3)** |
| **Unclear/not reported, n (%)** | **4 (7)** | **3 (12)** | **1 (3)** |
| **Met minimum criteria for infection** |  |  |  |
| Yes | 25 (41) | 11 (42) | 14 (40) |
| No | 23 (38) | 7 (27) | 16 (46) |
| Not applicable^2^ | 13 (21) | 8 (31) | 5 (14) |

*Denominator is total number of antimicrobial prescriptions. Antimicrobials may have greater than one documented indication.

† Indications outside of urinary, respiratory and skin/soft tissue infections.
